# HIV prevalence and associated factors among adolescent boys and young men in South Africa, 2017

**DOI:** 10.1101/2023.10.17.23297141

**Authors:** Tawanda Makusha, Musawenkosi Mabaso, Nompumelelo Zungu, Sizulu Moyo, Inbarani Naidoo, Sean Jooste, Karabo Mohapanele, Khangelani Zuma, Leickness Simbayi

## Abstract

**Introduction:** In South Africa, current epidemic control efforts include strategies to reach all young people and leave no one behind. Hence there is a need to track HIV among adolescent boys and young men (ABYM) to support these efforts. We examined HIV prevalence and associated factors among ABYM aged 15-24 years using the 2017 South African National HIV Prevalence, Incidence, Behaviour, and Communication Survey.

**Methods:** The cross-sectional survey used a multi-stage stratified cluster randomised sample. Descriptive statistics were used to characterise HIV prevalence among ABYM. Multivariate backward stepwise logistic regression was used to determine factors associated with HIV prevalence.

**Results:** A total of 4792 ABYM aged 15-24 years were interviewed of whom 4.01% [95% CI: 3.28-4.90] were HIV positive, translating to 255 366 ABYM living with HIV in the country in 2017. The odds of being HIV positive were significantly lower among ABYM with tertiary education level (AOR=0.06 [95% CI 0.01-0.50], p=0.009, employed (AOR=0.34 [95% CI: 0.14-0.81], p=0.015), and those who had tested and were aware of their HIV status (AOR=0.29 [95% CI: 0.10-0.83], p=0.015).

**Conclusion:** These findings suggest that ABYM with low educational attainment and those unemployed need to be reached with sexual and reproductive health interventions, including the promotion of uptake of HIV testing and awareness for this population group.

## Introduction

More than four decades into the epidemic, HIV continues to cause substantial illness and death among adolescents and young people in South Africa and other parts of the world due to low rates of HIV diagnosis, linkages to care, and initiation of and treatment adherence.^1–4^ In South Africa, in 2017, only 38.4% of adolescent girls and young women (AGYW) aged 15-19 years and 28.7% of boys and young men (ABYM) aged 15-19 years reported testing for HIV in the past 12 months and receiving the results of the last test.^5^ Globally, AGYW acquire HIV at significantly higher rates than ABYM. They have up to eight-fold higher rates of HIV infection compared to their male peers,^6–9^ with the gender divide especially pronounced in Eastern and Southern Africa.^10^ Focusing attention on HIV prevention and treatment among AGYW is therefore critical.

However, interventions and research focusing on the HIV service needs of ABYM, and strategies and interventions targeting this population group are notably absent; except for school-based interventions which are common to both sexes and voluntary medical male circumcision (VMMC).^11^ Opportunities are often missed to deliver facility-based HIV prevention messaging and counselling to infected and uninfected ABYM because most health services are often tailored for women and children. Funding support for HIV prevention for ABYM has also been limited compared to that for AGYW, resulting in insufficient data on the intersecting multi-level factors that contribute to HIV risk among them, and leading to limited evidence-based risk-reduction programmes.^10, 12^ Due to this neglect, there has been a failure to understand how gender disparities drive the burden of HIV and ill health for ABYM.

In addressing this gap, this paper examines the HIV prevalence and associated factors among ABYM aged 15-24 years using the 2017 South African National HIV Prevalence, Incidence, Behaviour, and Communication Survey. Although we know that HIV incidence and prevalence among men in sub-Saharan Africa peak at a much older age group than in women,^13^ ABYM are also at high risk of HIV infection. This is due to a combination of high-risk sexual behaviours including condomless sex, sex under the influence of alcohol, high partner turnover, early sexual debut, and multiple sexual partnerships as well as poor health-seeking behaviours, such as not getting tested for HIV and other sexually transmitted infections (STI) and poor levels linking to care among those who are HIV positive.^12, 14, 15^ Furthermore, the behavioural patterns formed in early adolescence are often ingrained in adulthood, setting the stage for ABYM’s increasing risk of HIV infection as they grow older.^16^ Early adolescence therefore provides opportunities for instilling positive sexual and reproductive health attitudes and behaviours.^10^

Previous studies in sub-Saharan Africa have associated education, marital status, employment, and income with HIV infection and prevalence. While a systematic review conducted in 2002 indicated a significant association between higher education attainment and a greater risk of HIV infection,^17^ subsequent studies have indicated that the patterns of association have been changing over time, despite considerable heterogeneity.^18, 19^ More studies show a protective effect of education, with higher educational attainment associated with decreased HIV risk.^20^ A study on HIV incidence in Uganda found that being enrolled in school was protective for both women and men.^13^ However, completing secondary school, relative to completing only primary school was protective only for men.

The same study found that marriage was associated with protection against HIV infection, with formerly married ABYM and AGYW having higher risks of new HIV acquisition compared to those who were currently married.^13^ Characteristics of partnerships may contribute to the risk of HIV acquisition. However, studies on age-disparate sexual partnerships and HIV risk have largely focused on AGYW and have produced mixed findings.^8, 21–23^ Little is known about the sexual partnerships of ABYM, including age-disparate sexual partnerships, although we know that multiple concurrent sexual partnerships are common.^24, 25^

A South African study on the trends and correlates of HIV prevalence among adolescents aged 12-19 years in different contexts and at various stages of the epidemic examined the association between risky sexual behaviours such as early sexual debut, inconsistent condom use, substance use (alcohol and drug use), peer pressure, and sensation-seeking behaviours and HIV prevalence.^26^ Results from this study indicated a statistically significant change in HIV prevalence among adolescent males over time. The HIV prevalence among adolescent males aged 12-19 years was estimated at 1.8% in 2008, declining to 1.6% in 2012 and then increasing markedly to 4.5% in 2017.^26^ Honing in on older adolescent males aged 15-19 years showed a steep increase in HIV prevalence between 2012 (0.7%) and 2017 (4.2%).^27^ There are substantial gaps in awareness of their HIV status among adolescents aged 10-19 years and reasons for this are complex.^27^ Makusha and colleagues (2022) found a significantly higher prevalence of HIV among males aged ≥15 years in South Africa who were unaware of their HIV status. Given this massive increase in HIV prevalence among ABYM and the limited evidence on associated factors in this generalised epidemic, this paper focuses on the HIV prevalence and associated factors among ABYM in South Africa.

## Methods

### Study data and sample

This secondary analysis used data from the 2017 South African National HIV Prevalence, Incidence, Behaviour, and Communication Survey, a nationally representative household survey described in detail elsewhere.^28, 29^ Briefly, participants were selected using multi-stage stratified cluster sampling. A systematic probability sample of 15 households was drawn from each of 1000 small area layers (SALs) selected randomly, probability proportional to size, from a master sample stratified by province, locality type (urban, rural formal, and rural informal/tribal areas), and predominant race groups in urban areas obtained from an updated database from Statistics South Africa.^30^ A detailed head of the household questionnaire and three age-specific questionnaires were used to solicit information about socio-demographic factors and information about HIV knowledge, attitudes, practices, and behaviours of consenting participants. All members of the selected households were invited to participate in the survey.

Dried blood spots (DBS) specimens were collected from consenting participants and tested anonymously for HIV antibodies using a testing algorithm with three different enzyme immunoassays (EIAs). All samples that were HIV positive on the first two EIAs (Roche Elecys HIV Ag/Ab assay, Roche Diagnostics, Mannheim, Germany, and Gene screen Ultra HIV Ag/Ab assay, Bio-Rad Laboratories, Hercules, CA, USA) were subjected to a nucleic acid amplification test (COBAS AmpliPrep/Cobas Taqman HIV-1 Qualitative Test, v2.0, Roche Molecular Systems, Branchburg, NJ, USA) for the final determination of HIV status.

The data was benchmarked to the mid-year estimates for 2017 to generalize the findings to the South African population.^30^ The current analysis is based on a sub-sample of ABYM aged 15-24 years who tested for HIV.

## Measures

### Dependant variable

The primary outcome variable was HIV serostatus (HIV positive =1 and HIV negative = 0) among ABYM aged 15 to 24 years.

### Independent variables

Socio-demographic variables were age, race (Black African and other race groups), educational level (no education/primary, secondary and tertiary), employment status (unemployed and employed), asset-based socio-economic status (SES) representing a continuum of household SES from the poorest (lowest quintile) to the least poor (highest quintile), locality type (urban areas, rural informal/tribal, rural formal/farm areas) and nine South African provinces. Socio-behavioural variables were risky sexual behaviours, including age at early sexual debut (less than 15 years of age, and more than 15 years of age), age-disparate sexual partnerships (within 5 years age, younger than 5 years and older than 5 years), number of sexual partners in the last 12 months (one partner, and two or more sexual partners), condom use at last sex (no and yes), consistent condom use (no and yes). Alcohol use risk score (abstainers, low risk drinkers, high risk drinkers and hazardous risk drinkers) based on the Alcohol Use Disorder Identification Test (AUDIT) scale.^31, 32^ HIV-related factors such as correct HIV knowledge and myth rejection (no and yes), self-perceived risk of acquiring HIV (low and high), testing for HIV, and awareness of HIV status (no and yes) were also included.

### Statistical analysis

All statistical analyses were performed using Stata software version 15.0 (Stata Corp, College Station, Texas, USA) using weights to adjust for the complex survey design. Descriptive statistics were used to summarise socio-demographic, socio-behavioural, and HIV-related factors, including HIV prevalence. Differences between categorical variables were assessed using the chi-square test. A multivariate stepwise backward logistic regression selection method was used to determine factors associated with HIV prevalence. Adjusted odds ratios (aORs) with 95% confidence intervals (CI) and a p-value < 0.05 were used to ascertain statistical significance.

### Ethical approval

The survey protocol was approved by the Human Sciences Research Council’s Research Ethics Committee (REC: 4/18/11/15) and the Associate Director for Science, Center for Global Health, Centers for Disease Control and Prevention (CDC), GA, USA. Informed consent was sought from all participants aged 18 years and older. However, informed consent was first sought from parents/guardians of participants aged 15–17 years, and then assent was obtained from the youth themselves.

## Results

### Sample characteristics

The study sample consisted of 4792 ABYM aged 15-24 years. About half were aged 20-24 years. Most were Black African, unmarried, had secondary-level education and were unemployed. About half of the sample was from low and high-SES households, and most were from urban areas as indicated in Table 1.

**Table 1:** Socio-demographic characteristics, ABYM aged 15-24 years, South Africa 2017.

| **Variables** | **N** | **%** |
| --- | --- | --- |
| **Age in years** |  |  |
| 15-19 | 1932 | 47.8 |
| 20-24 | 1612 | 52.2 |
| **Race groups** |  |  |
| Black African | 4035 | 83.4 |
| Other | 757 | 16.6 |
| **Marital status** |  |  |
| Married | 18 | 0.5 |
| Never married | 3328 | 99.5 |
| **Educational level** |  |  |
| No education/Primary | 147 | 9.1 |
| Secondary | 1141 | 81.4 |
| Tertiary | 102 | 9.5 |
| **Employment status** |  |  |
| Unemployed | 2899 | 87.5 |
| Employed | 423 | 12.5 |
| **Asset-based SES** |  |  |
| Low SES | 2195 | 46.7 |
| High SES | 2052 | 53.3 |
| **Locality type** |  |  |
| Urban areas | 2292 | 60.1 |
| Rural informal/tribal areas | 1995 | 34.9 |
| Rural formal/farm areas | 505 | 5.0 |

Sub-totals do not all add to the sample total due to non-response and/or missing data, CI-confidence intervals, and socioeconomic status. Other race groups refer to White, Coloured or mixed race and Asian/Indian origin combined.

Table 2 shows that most participants had a sexual debut at age 15 years and older, had sexual partners within a five-year age gap of their age group, had one sexual partner in the last 12 months, used a condom in the last sex act, did not use condoms consistently, abstained from alcohol, did not have the correct HIV knowledge and myth rejection, had a low self-perceived risk of HIV, and had tested for HIV and were aware of their status.

**Table 2:** HIV-related factors among ABYM aged 15-24 years, South Africa 2017.

| **Variables** | **N** | **%** |
| --- | --- | --- |
| **Age of sexual debut** |  |  |
| younger than 15 years | 324 | 10.2 |
| 15 years and older | 3205 | 89.8 |
| **Age disparate relationships** |  |  |
| Within 5 years | 1210 | 90.4 |
| Younger than 5 years | 87 | 6.1 |
| Older than 5 years | 45 | 3.5 |
| **Number of sexual partners in the last 12 months** |  |  |
| One partner | 1009 | 72.4 |
| Two or more partners | 345 | 27.6 |
| **Condom use at last sex act** |  |  |
| No | 440 | 32.2 |
| Yes | 916 | 67.8 |
| **Consistent condom use** |  |  |
| No | 1295 | 95.3 |
| Yes | 54 | 4.7 |
| **Alcohol AUDIT score** |  |  |
| Abstainers | 3338 | 75.2 |
| Low drinkers (1-7) | 553 | 15.5 |
| High-risk drinkers (8-19) | 287 | 8.2 |
| Hazardous drinkers (20+) | 37 | 1.1 |
| **Correct HIV knowledge and myth rejection** |  |  |
| No | 3123 | 69.0 |
| Yes | 1402 | 31.0 |
| **Self-perceived risk of acquiring HIV** |  |  |
| Low | 3839 | 86.2 |
| High | 572 | 13.8 |
| **Tested and awareness of HIV status** |  |  |
| No | 3012 | 68.2 |
| Yes | 1396 | 31.8 |

Sub-totals do not all add to the total due to non-response and/or missing data, CI-confidence intervals, SES-socioeconomic status, and AUDIT score – Alcohol Use Disorder Identification Test.

### HIV prevalence and sample characteristics

Overall HIV prevalence was 4.2% (95% CI 3.0-5.8) among ABYM aged 15 years and older, translating to a total of 255 366 ABYM living with HIV in the country in 2017.

Figure 1 shows that HIV prevalence among ABYM was significantly higher (p= 0.016) in Mpumalanga (8.5%, 95% CI: 5.8-12.3), followed by KwaZulu-Natal (5.5%, 95% CI: 4.1-7.5).

**Figure 1:**
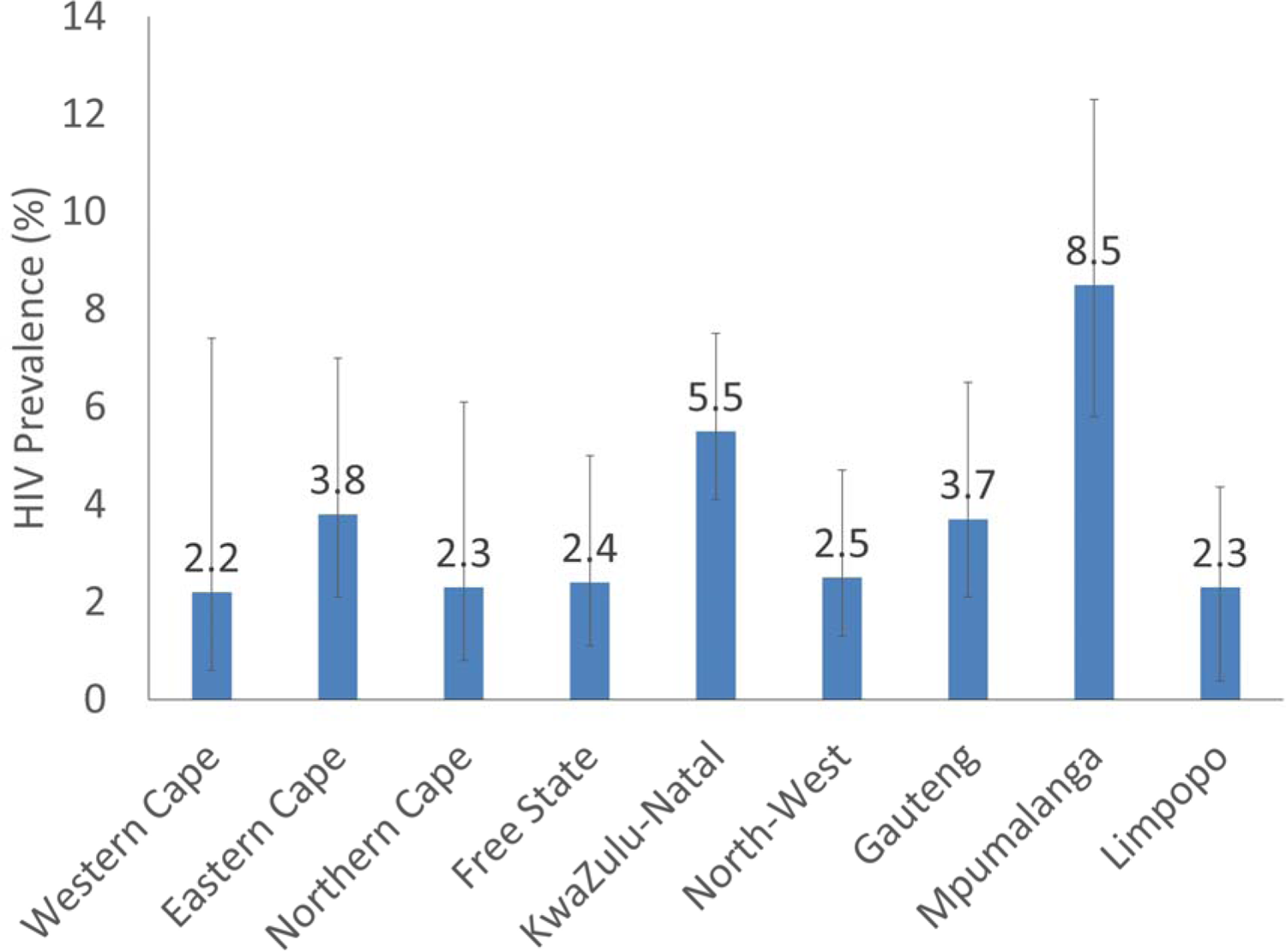
HIV prevalence among ABYM by the provinces in South Africa, 2017

Table 3 shows that HIV prevalence among ABYM was significantly higher among Black Africans (4.6% [95% CI: 3.7-5.6], p=0.003), those with no education/primary-level education (8.0% [95% CI: 3.9-15.5], p=0.043), those residing in rural formal/farm areas (9.0%, [95% CI: 6.1-12.9], p= 0.005).

**Table 3:** HIV prevalence among ABYM aged 15-24 years by socio-demographic characteristics, South Africa 2017.

| Variables | N | (%) | 95% CI | p-value |
| --- | --- | --- | --- | --- |
| **Age in years** |  |  |  | 0.240 |
| 15-19 | 1932 | 4.2 | 3.0-5.8 |  |
| 20-24 | 1612 | 5.4 | 4.1-7.1 |  |
| **Race group** |  |  |  | 0.003 |
| Black African | 4035 | 4.6 | 3.7-5.6 |  |
| Other | 757 | 1.1 | 0.4-3.0 |  |
| **Educational level** |  |  |  | 0.043 |
| No education/Primary school | 147 | 8.0 | 3.9-15.5 |  |
| Secondary school | 1141 | 7.1 | 4.9-10.1 |  |
| Tertiary | 102 | 1.3 | 0.5-3.3 |  |
| **Employment status** |  |  |  | 0.588 |
| Unemployed | 2899 | 5.0 | 3.9-6.3 |  |
| Employed | 423 | 4.3 | 2.6-7.0 |  |
| **Asset-based SES** |  |  |  | 0.152 |
| Low SES | 2195 | 4.7 | 3.6-6.0 |  |
| High SES | 2052 | 3.4 | 2.3-4.9 |  |
| **Locality type** |  |  |  | 0.005 |
| Urban areas | 2292 | 3.7 | 2.7-5.1 |  |
| Rural informal/tribal areas | 1995 | 3.8 | 2.9-5.0 |  |
| Rural formal/farm areas | 505 | 9.0 | 6.1-12.9 |  |

Sub-totals do not all add to the total due to non-response and/or missing data, CI-confidence intervals, and SES-socioeconomic status.

Table 4 shows no statistically significant association between HIV prevalence and various socio-behavioural characteristics and HIV-related factors among ABYM. However, HIV prevalence was higher among those who had a sexual debut at younger than 15 years, those who reported having a sexual partner five years younger than their age, those who reported having one sexual partner, those who reported no condom use at last sex act, hazardous drinkers and those who were tested and aware of their HIV status though not statistically significant.

**Table 4:** HIV prevalence among ABYM aged 15-24 years by socio-behavioural characteristics and HIV-related factors, South Africa 2017.

| Variables | N | (%) | 95% CI | p-value |
| --- | --- | --- | --- | --- |
| **Age of sexual debut** |  |  |  | 0.579 |
| Less than 15 years | 324 | 6.1 | 2.4-14.4 |  |
| 15 years and older | 3205 | 4.7 | 3.8-5.8 |  |
| **Age disparate relationships** |  |  |  | 0.667 |
| Within 5 years | 1210 | 4.4 | 2.9-6.5 |  |
| Younger than 5 years | 87 | 6.5 | 2.6-15.2 |  |
| Older than 5 years | 45 | 6.3 | 1.4-24.1 |  |
| **Number of sexual partners in the last 12 months** |  |  |  | 0.096 |
| One partner | 1009 | 5.2 | 3.4-7.9 |  |
| Two or more partners | 345 | 2.5 | 1.1-5.6 |  |
| **Condom use at the last sex act** |  |  |  | 0.151 |
| No | 440 | 6.4 | 3.1-12.7 |  |
| Yes | 916 | 3.5 | 2.3-5.2 |  |
| **Consistent condom use** |  |  |  | 0.928 |
| No | 1295 | 4.4 | 2.9-6.5 |  |
| Yes | 54 | 4.0 | 0.8-18.2 |  |
| **Alcohol AUDIT score** |  |  |  | 0.775 |
| Abstainers | 3338 | 3.9 | 3.0-4.9 |  |
| Low-risk drinkers (1-7) | 553 | 4.5 | 2.7-7.4 |  |
| High-risk drinkers (8-19) | 287 | 5.1 | 2.4-10.5 |  |
| Hazardous drinkers (20+) | 37 | 7.0 | 1.0-36.3 |  |
| **Correct HIV knowledge and myth rejection** |  |  |  | 0.213 |
| No | 3123 | 4.4 | 3.4-5.6 |  |
| Yes | 1402 | 3.3 | 2.2-4.9 |  |
| **Self-perceived risk of HIV** |  |  |  | 0.929 |
| Low | 3839 | 3.4 | 2.6-4.5 |  |
| High | 572 | 3.5 | 2.2-5.5 |  |
| **Tested and awareness of HIV status** |  |  |  | 0.664 |
| No | 3012 | 4.0 | 3.0-5.4 |  |
| Yes | 1396 | 3.7 | 2.6-5.1 |  |

Sub-totals do not all add to the total due to non-response and/or missing data, CI-confidence intervals, SES-socioeconomic status, and AUDIT score – Alcohol Use Disorder Identification Test.

### Factors associated with HIV prevalence among ABYM aged 15-24 years

In the final multivariate model (Figure 2), the odds of being HIV positive were significantly lower among ABYM with tertiary education level than those with no education/primary school level education (aOR=0.06 [95% CI 0.01-0.50], p=0.009). The odds were also significantly lower among ABYM who were employed (aOR=0.34 [95% CI: 0.14-0.81], p=0.015), and among those who indicated that they had tested and were aware of their HIV status (AOR=0.29 [95% CI: 0.10-0.83], p=0.015).

**Figure 2:**
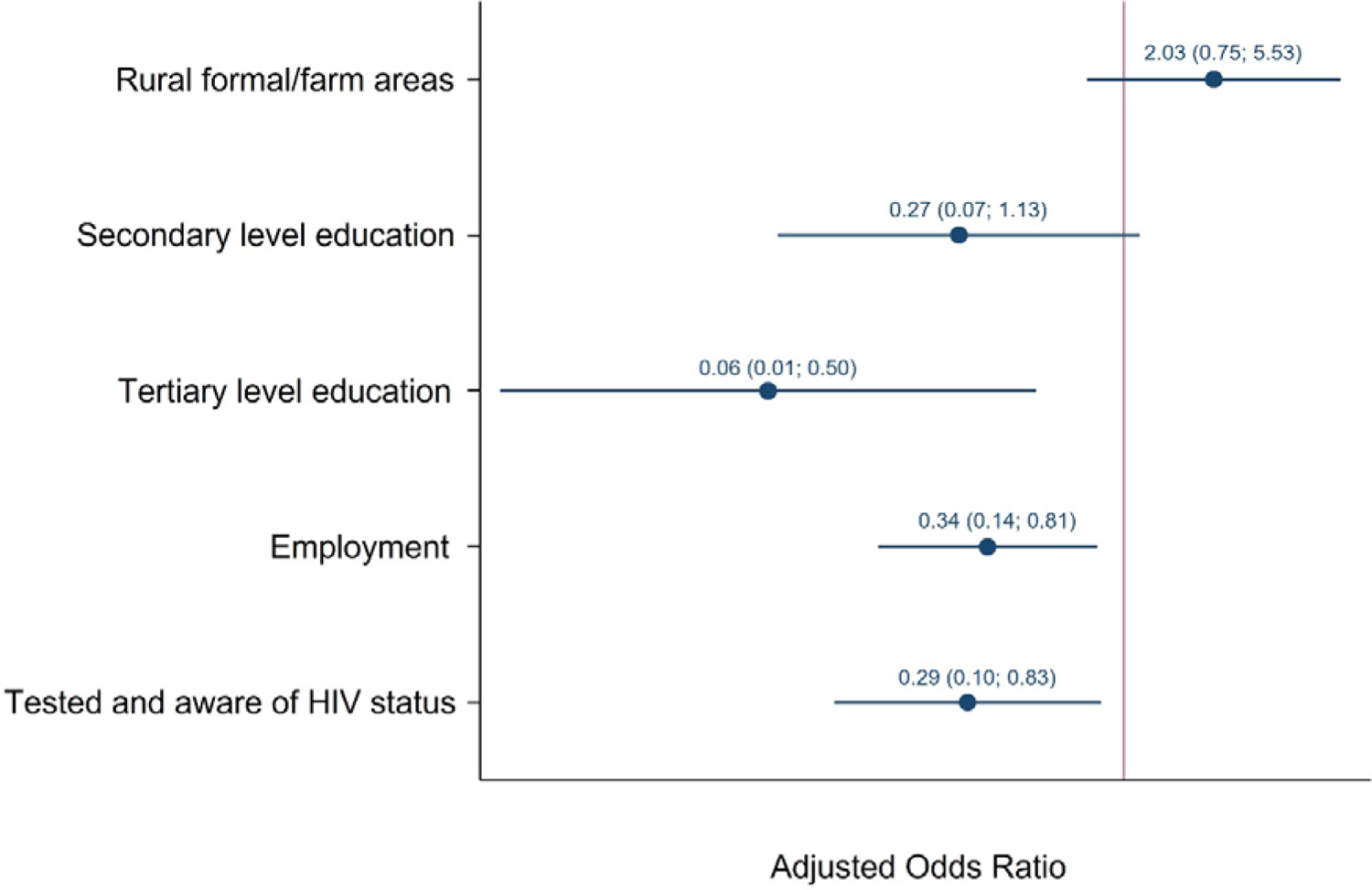
Coefficient plot showing the model of factors associated with HIV prevalence among ABYM aged 15-24 years, South Africa 2017

## Discussion

This is the first nationally representative study focusing on the HIV prevalence and associated factors among ABYM aged 15-24 years in a generalised epidemic. The results revealed an HIV prevalence of 4.01% among ABYM aged 15-24 years. This translates to just over a quarter of a million ABYM living with HIV in 2017 in the country, highlighting the HIV burden in this population group. This is substantial since HIV incidence in this population group increased between 2012 and 2017, indicating an urgent need for evidence-based HIV programmes focused on ABYM in South Africa.

The results highlight a significant association between HIV prevalence and selected socio-demographic factors. HIV prevalence was significantly higher among ABYM residing in Mpumalanga and KwaZulu-Natal provinces, among Black Africans, those from rural formal/farm areas, and among those with no education or with primary-level education. Overall, these results reflect the state of the South African HIV epidemic, with high HIV prevalence in rural provinces, and the concentration of infections mainly among Black Africans residing in areas of widespread poverty, unemployment, and lack of quality education.^8, 33^ These structural conditions contribute to increased vulnerability and risk of HIV acquisition among Black African ABYM in South Africa.

The final multivariate model results suggest that HIV prevalence was significantly associated with those ABYM with no education or low educational attainment, those who were unemployed, and those who tested and were unaware of their HIV status. This is consistent with our previous analyses among adolescents aged 12-19 years where we found that adolescents who did not attend an education institution and who were unemployed were more likely to be HIV positive.^26^ These observations likely reflect socio-economic disparities that increase the risk of HIV infection among impoverished individuals.

Studies have shown that educational play a significant role in fostering safe sexual behaviours among men in a generalised epidemics. Such behaviours include having fewer sexual partners, increased use of condoms, increased utilization of sexual and reproductive health services, and better infection prevention.^18, 19^ According to Makusha and colleagues (2022), ensuring adolescents complete both school and higher education has multiple benefits. These include economic empowerment and exposure to HIV-related knowledge and information on risk-reducing behaviours through educational institution-based interventions.

Furthermore, studies have shown a significant link between unemployment and the prevalence of HIV. It has been suggested that unemployment can result in young men engaging in risky sexual behavior, which increases their risk of contracting HIV (Baral et al., 2011). Programs promoting wellness in the workplace can provide knowledge and information about HIV, empowering individuals to lead healthier lives. Employment also provides financial independence, allowing individuals to access education and healthcare, which can increase awareness and lower vulnerability to HIV.^34^ Therefore, initiatives promoting sustained youth employment can help mitigate the impact of poverty and HIV on young people.

The findings suggest that adolescent boys and young men (ABYM) who were unaware of their HIV status and had not undergone testing are at a higher risk of contracting HIV. In line with the data for the general population in South Africa, despite an increase in awareness of HIV status, most HIV-positive ABYM are still unaware of their serological status and thus do not receive appropriate treatment. ^28, 35, 36^ These findings are also consistent with studies conducted in Uganda, which demonstrated that self-assessment of HIV risk was strongly linked to HIV acquisition.^13^ These observations emphasize the importance of expanding HIV testing and counselling services to help ABYM become aware of their HIV status and prevent transmission to themselves and others.

### Limitations

This study used self-report and given the sensitive nature of questions related sexual behaviours may have induced recall and social desirability bias. The causal relationships between HIV prevalence and selected covariates could not be deduced due to cross-sectional nature of the survey which only allows for assessment of associations. The secondary analysis may also be limited by unmeasured or unobserved covariates. However, given that the study is nationally representative, the current findings can be generalized to ABYM aged 15-24 years in the country.

## Conclusion

The study highlights the need for HIV interventions tailored for out of school ABYM, and those with low educational attainment. This should also include efforts to keep AYBM in school and ensure that remain to attain higher education. These findings also suggest that employment might play a role in reducing HIV infection. This supports initiative for creating income generating activities for ABYM to reduce drivers of the HIV epidemic in this population group. However, research is needed to understand the mechanisms through which employment impacts HIV. Finally, the findings suggest a need for increasing the uptake of HIV testing and awareness in the study population. This is essential since testing is the gateway to treatment and effective treatment is a great HIV prevention tool.

## Data Availability

All data and related metadata underlying the findings reported in this submitted
manuscript are deposited in an appropriate public repository:
https://ghdx.healthdata.org/series/south-africa-national-hiv-prevalence-incidence-behavior-and-communication-survey-sabssm

